# Intracranial pressure and pulsatility in different head and body positions

**DOI:** 10.1101/2024.06.20.24309240

**Authors:** MJ Bancroft, EM Moncur, AL Peters, L D’Antona, L Thorne, LD Watkins, BL Day, AK Toma

## Abstract

Intracranial pressure (ICP) is typically measured with the head in a neutral position whilst the body is in an upright or supine posture. The effect of body position on ICP is well studied, with ICP greater when supine than when upright. In daily life the head is frequently moved away from the neutral position but how this impacts ICP dynamics is unclear. Knowledge of ICP dynamics in different head-on-body positions may improve future treatments that restore normal ICP dynamics such as cerebrospinal fluid (CSF) drainage shunts.

We recruited 57 relatively well, ambulatory patients undergoing clinical ICP monitoring for investigation of possible CSF dynamics disturbances. Forty-one patients were non-shunted, seven had a working shunt and nine had a malfunctioning shunt. We measured ICP and ICP pulsatility (pulse amplitude) over 10 or 20s in different combinations of head and body positions. Positions included right and left head turn and forward tilt in upright (seated, standing) and supine body positions, and right and left lateral tilt and backward tilt in upright body positions.

ICP increased by 3-9 mmHg, on average, when the head moved away from neutral to each head position in upright and supine body positions, except for head forward tilt when supine, where ICP did not change. The increase in ICP with head turn and forward tilt in upright body positions was larger in patients with a malfunctioning shunt than with no shunt or a functioning shunt. Pulsatility also increased by 0.5-2 mmHg on average when the head moved away from neutral to each head position in upright and supine body positions, except for head forward tilt in upright body positions where pulsatility slightly decreased by 0.7 mmHg on average.

ICP and pulsatility generally increase when the head is moved away from the neutral position, but this depends on a combination of head and body position and shunt status. We propose our results can be explained by a combination of changes to neck vasculature and head orientation relative to gravity. Our findings provide potential reason for patient reports that ICP-related symptoms can be induced and/or exacerbated by head movement and could explain behaviours that avoid excess head movement, such as turning the body rather than the head when looking to the side.

## INTRODUCTION

Brain health depends critically on the regulation of intracranial pressure (ICP). ICP reflects the force exerted within the skull by the volumes of the brain, cerebrospinal fluid (CSF) and intracerebral blood^1,2^, and is in a constant state of flux as fluid flows into and drains from the cranial compartment. Individuals with abnormal ICP dynamics experience wide-ranging symptoms including vision impairment, headache, and gait and balance disturbances.^3,4^ CSF drainage shunts are commonly used to treat hydrocephalus, but current shunt technologies are limited by relatively high failure rates and a lack of adequate knowledge about the pathophysiology of ICP with which to guide optimal shunt settings.^5–7^ Further understanding of ICP dynamics is crucial to allow technological advances that could transform restorative treatments such as the proposed ‘smart’ CSF-diverting shunt.^8^

Current knowledge of ICP dynamics is mostly from studies of patients in neurocritical care when lying supine, with fewer studies of ambulatory individuals under other circumstances. One accepted determinant of ICP is body position. ICP is higher in horizontal body positions such as when lying supine than in upright body positions like sitting or standing.^9–13^ The dependence of ICP on body position reflects the influence of gravity on fluid shifts within the craniospinal compartment. When in an upright body position, fluid is shifted away from the head by gravity, but as the head and body is tilted further from upright fluid is redistributed towards the head and ICP increases.^14–16^

Head position can be changed independently of body position in daily life, such as when turning the head to the side. How the position of the head on the body influences ICP is underexplored. Positioning the head away from the neutral position whilst body position is maintained can increase ICP in neurocritical care patients^17–23^, possibly due to torsion or compression of neck vasculature restricting outflow of fluid from the cranial compartment.^19,24–30^ Despite this, most studies either do not report head-on-body position or do not manipulate head-on-body position, instead manipulating head and body position together such as during tilt table testing.

It is unclear whether data from individuals in neurocritical care is applicable to comparatively well and ambulatory individuals such as those undergoing elective ICP monitoring for diagnosis and treatment of idiopathic intracranial hypertension (IIH), Chiari malformation, hydrocephalus or other CSF-related disturbances. One key difference is that studies in neurocritical care individuals inevitably ‘passively’ position the head, where the head is moved into position *for* the patient rather than actively moved *by* the patient.^22,23^ Passive positioning is rare in daily life for most humans and active positioning may better represent ICP dynamics^31^, particularly given recent evidence that suboccipital muscle-driven head movement may modulate CSF circulation.^32–34^

Whether the same head-on-body position influences ICP identically across body positions is also underexplored. Consider the example of neck flexion, where the head is tilted forward such that the chin is moved towards the chest and held in position. When in an upright body position this head position will compress the neck and move the head from an upright towards a horizontal position. Both factors could be expected to increase ICP independently. However, when in a supine body position the same head movement may have different implications for ICP. The neck will be compressed in a similar manner as when upright, and could still be expected to increase ICP, but the head is moved from a horizontal towards an upright position, which is expected to decrease ICP. The two factors are now opposed but whether an increase, decrease, or no change in ICP occurs is unclear.

ICP pulsatility (pulse amplitude) is the amplitude of the cardiac-induced fluctuation in ICP and is an important, indirect biomarker of intracranial compliance.^35–37^ Compliance is the capacity of the intracranial compartment to buffer an increase in volume against an increase in pressure.^36,37^ Pulsatility and intracranial compliance are inversely related such that greater pulsatility reflects decreased compliance. To our knowledge, pulsatility has not yet been reported in different head positions in non-neurocritical care individuals.^38^

Here, we measure intraparenchymal ICP and pulsatility, plus head and body position, in a relatively well, ambulatory cohort of mixed age, body habitus, pathology, and shunt status. Patients adopted various head positions for up to 20s whilst either seated, standing, or supine. Head positions spanned commonly adopted postures, with head movement in different directions, about different axes of motion, and resulting in different changes in head orientation relative to gravity.

## MATERIALS AND METHODS

The study conformed to the *Declaration of Helsinki* and ethical approval was granted by local Research Ethics Committee (project ID 15/0769). All participants provided written informed consent to clinical ICP monitoring and involvement in the study, separately.

### Patients

We recruited 57 patients who were undergoing elective ICP monitoring at our centre as part of clinically indicated assessment of possible CSF dynamics disturbances (National Hospital for Neurology and Neurosurgery, London). Table 1 summarises the patients’ characteristics, clinical indications for ICP monitoring, and diagnosed pathologies. Sixteen patients had a shunt in situ at the time of monitoring. Suspected shunt malfunction was the clinical indication for ICP monitoring in all 16 shunted patients. Of these patients, nine were subsequently diagnosed with a malfunctioning shunt and seven to have a functioning shunt. This diagnosis was made by a multi-disciplinary team based on clinical and radiological findings in combination with ICP monitoring results.

**Table 1.**
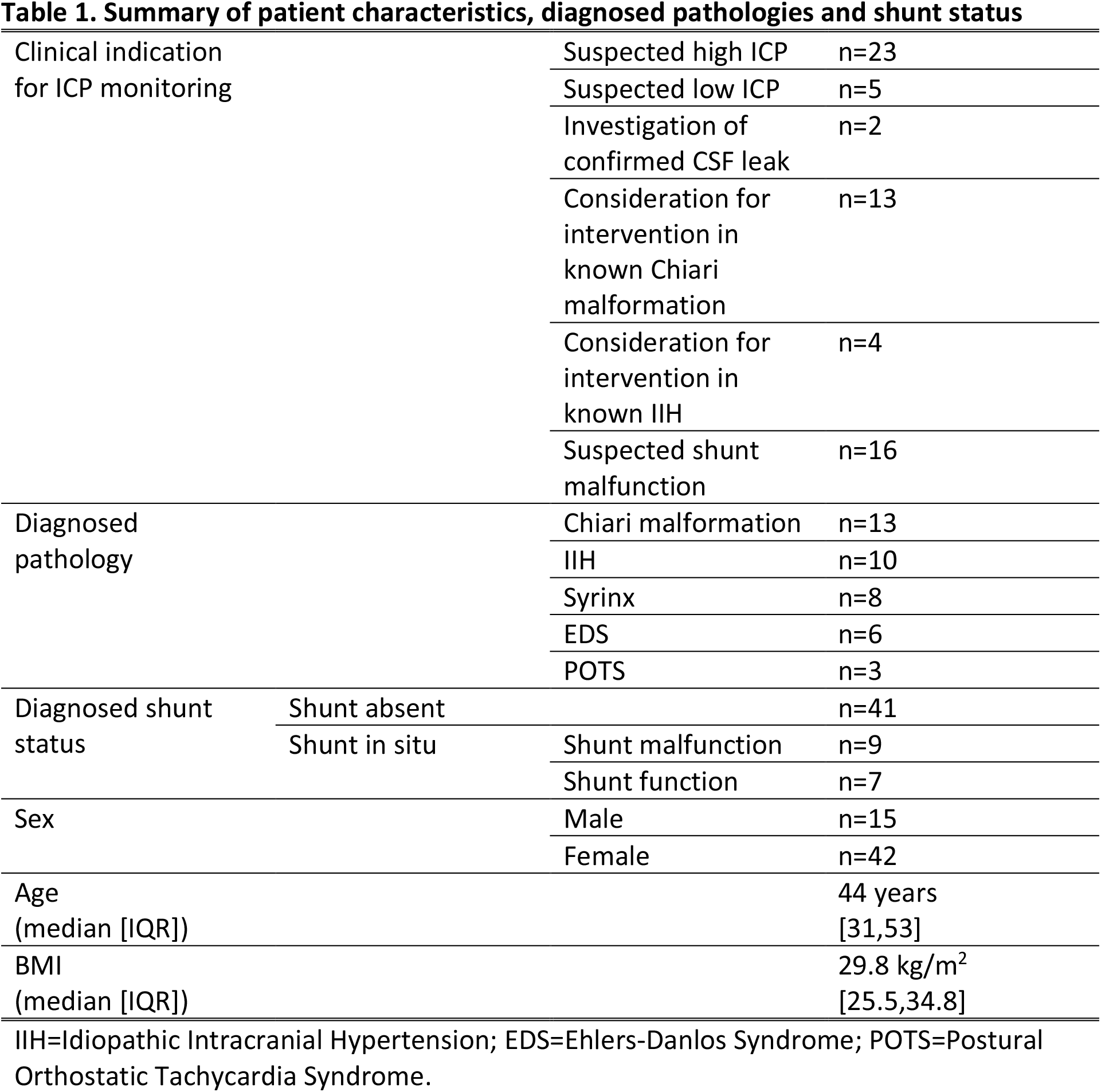
Summary of patient characteristics, diagnosed pathologies and shunt status.

### Protocol

Patients completed a short battery of recordings where they adopted a sequence of different head positions while in different body positions (seated, standing, or supine). The head and body positions were intended both to reflect postures commonly adopted in life and to encompass positions involving movement about each of the three cardinal axes of head angular motion (yaw, pitch, roll). Head positions were: (1) right or left yaw, describing neck rotation to point the nose over the shoulder, which we term head *turn*; (2) forward or backward pitch, describing neck flexion or extension to move the chin to the chest or nose to point upward, respectively, which we term head *tilt*; and (3) right or left roll, describing neck lateral flexion to move the ear toward the ipsilateral shoulder, which we term head *lateral tilt*. Each head position is depicted in Figs. 1-5.

**Figure 1.**
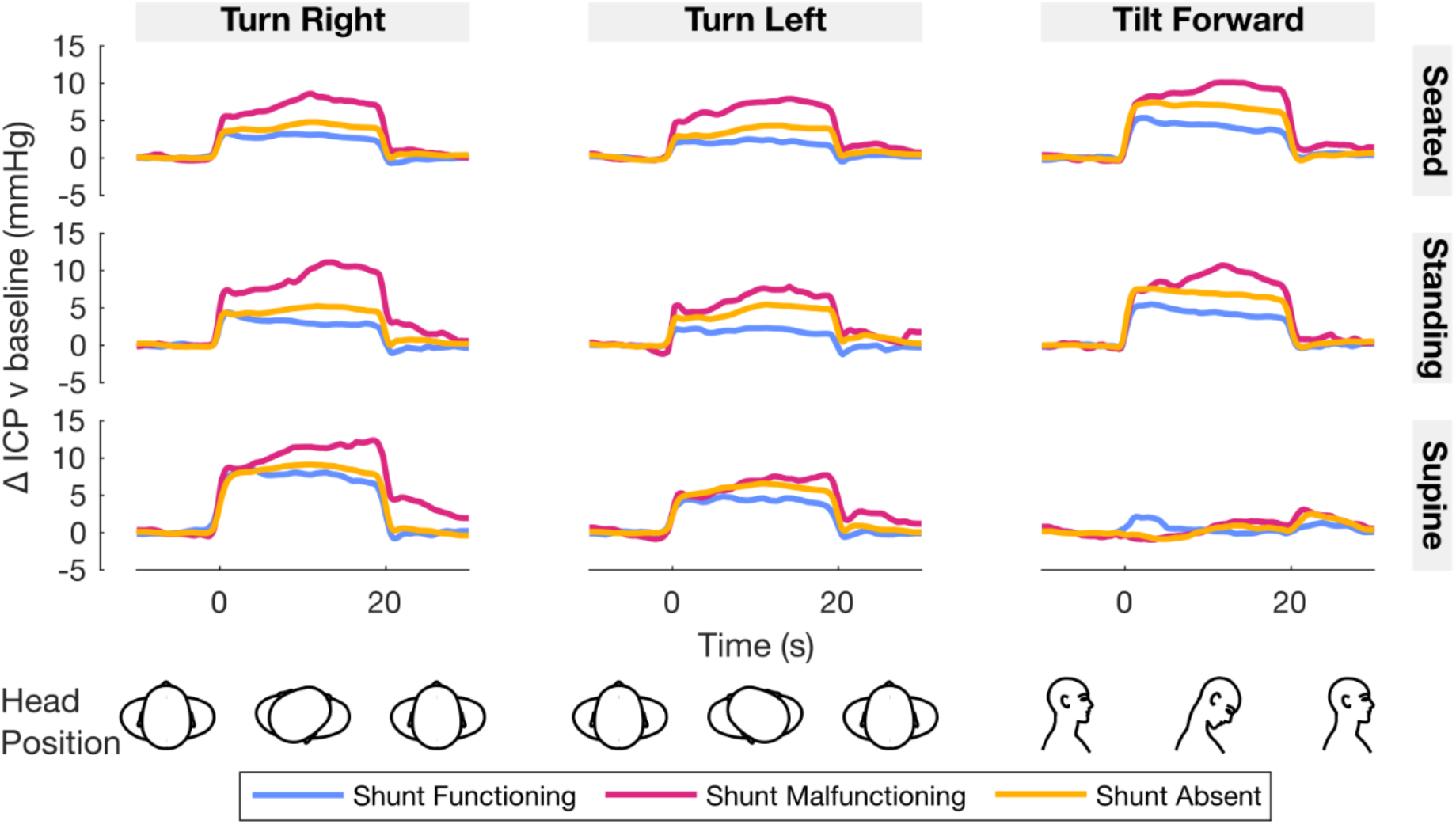
Effect of head turn and forward tilt on ICP over time. Traces are the mean change in ICP over time relative to baseline (head neutral) for each head position (left column: turn right; middle column: turn left; right column: tilt forward) in each body position (top row: seated; middle row: standing; bottom row: supine) and shunt status group (blue: shunt functioning; magenta: shunt malfunctioning; yellow: shunt absent). The head was moved from neutral into each position at time=0s and back to neutral at time=20s. For plotting, ICP was averaged from pulse-to-pulse to remove pulsatile variation prior to averaging per individual and group. Graphics at bottom of figure aid interpretation of head positions and timings.

Patients stood in the anatomical position at the beginning of each recording and then transferred to either a seated, standing, or supine body position and adopted a neutral head posture (nose pointing forward approximately perpendicular to the chest). Head position sequences were initiated after 60 s of rest in each body position to allow stabilisation of ICP. Left and right head turn, and forward tilt positions were measured in seated, standing, and supine body positions. Left and right lateral tilt was measured in the seated and standing body positions and backward tilt was measured in the seated body position only as they were viewed to be atypical postures to adopt when supine.

The patient maintained each head position for 20 s before moving their head back to the neutral position for 20 s, after which the next head position was adopted. Backward tilt was maintained for 10 s owing to prior clinical concern that 20 s in this head position could cause discomfort.

Head position sequences were performed in a set order to aid task compliance: turn right then turn left then forward tilt whilst seated, standing, or supine; lateral tilt right then lateral tilt left whilst seated or standing. Three repeats of each head position sequence were measured in each body position. Backward tilt whilst seated was performed in isolation after completion of all seated right/left turn and forward tilt repeats. The head rested on one low-profile pillow whilst supine.

Two patients with shunt malfunction completed head turn and forward tilt positions in only two of the three body positions (one patient did not do standing, the other did not do supine). The remaining 55 patients completed head turn and forward tilt positions in each body position. A subset of patients completed lateral tilt (n=39) and backward tilt (n=21) head positions owing to a protocol update.

### Intracranial pressure measurement

ICP was measured by wire probe surgically inserted into the frontal lobe parenchyma via an incision posterior to the hairline (Neurovent-P intraparenchymal ICP bolt, Raumedic). The ICP probe was inserted into the right frontal lobe unless clinical indication necessitated a left-sided insertion (n=2). The ICP probe insertion procedure used at our centre has been described previously.^39^ Raw ICP data were sampled at 100 Hz and recorded on a storage device (MPR1, Raumedic).

### Head and body position measurement

Head and body position were sampled at 100 Hz by wireless inertial measurement units (IMU) placed on the forehead and sternum (Xsens MTw Awinda, Netherlands).^40^ IMUs were attached to the patient via elasticated non-slip bands strapped around the head and chest. Each IMU contained 3D accelerometers, gyroscopes and magnetometers.

ICP and IMU data were collected simultaneously on separate devices. We temporally synchronised the data for offline analysis by sending a voltage from the IMU to the ICP storage device at the start and end of each recording.

### Data analysis

IMU data was collected in MT Manager v4.6^41^ and exported via Xsens’ proprietary strap-down integration and Kalman filter algorithms, yielding 3D head (forehead) and body (sternum) angular velocity and orientation relative to the world frame per sample.^40^ We used a static sensor-to-segment calibration procedure to compute head orientation relative to body orientation (herein termed head-on-body angle that technically refers to head-on-body angular displacement relative to head neutral).^42–44^ First, the IMU axis was aligned with the segment axis whilst the participant was in the anatomical position at the start of each recording; second, head relative to body orientation was computed per sample by matrix multiplication of the head-aligned rotation matrix by the transposed body-aligned rotation matrix; third, head relative to body orientation was offset by the average orientation in the 60 s period of rest before initiation of each head position sequence to return head-on-body angle relative to head neutral. The magnitude of the head-on-body angle relative to the neutral position was represented in axis-angle format to allow a common measure across directions and planes of motion. Head-on-body angle and head angle relative to vertical in each head and body position are summarised in Supplementary Fig 1. and Supplementary Table 1.

Periods in each head position were parsed from periods of movement between head positions using head-on-body angular velocity. Periods of movement between head positions were discarded for analyses, leaving only periods where the head was maintained in each position. Head and body position were labelled based on head-on-body angle and body orientation using an interactive Matlab script.

ICP was smoothed by a dual-pass (zero lag), 4^th^ order Butterworth low-pass filter with a 10 Hz cut off. Pulsatility was computed using a custom-written Matlab algorithm that calculated the difference in ICP (i.e. amplitude) between successive diastolic and systolic peaks in continuous ICP data within the heart-rate bandwidth. ICP, pulsatility and head-on-body angle were median averaged over each period in each head position, and then mean averaged across repetitions for each participant. To examine the effect of head position within each body position, we then subtracted the baseline (head neutral position) value from each head position value. We reasoned that an effect of head-on-body position would be evidenced by a change from baseline statistically different from zero.

Data was analysed using custom-written code in Matlab 2021a (Mathworks). Head-on-body angle data were averaged using appropriate circular statistical methods.^45^

### Statistical analysis

Data was statistically analysed by multi-level mixed-effects linear regression models in Stata v18 (StataCorp LLC, College Station, TX). We initially examined whether ICP and pulsatility in the head neutral position varied by body position, shunt status and diagnosed pathology (Table 1). We then examined the effect of head position on ICP and pulsatility relative to the head neutral position (baseline; see ‘Data analysis’). Separate models were fitted for backward tilt and lateral tilt data due to these head positions being recorded only in a subset of patients and in upright body positions, and over 10 s rather than 20 s for backward tilt (see ‘Protocol’). Terms were iteratively added to the models and included if likelihood ratio testing suggested an improved model fit. In the absence of prior data to inform specific hypotheses about effects of head-on-body position in each diagnosed pathology or shunt status group, we instead tested whether each combination improved model fit but used an approximately Šidák-adjusted alpha level (p<0.001) for likelihood ratio testing. We tested for effects of: head position, body position, head position by body position interaction, shunt status and its interaction by head and body position, each diagnosed pathology and its interaction by head and body position, baseline ICP, baseline pulsatility, age, sex, BMI, hypertension, and head-on-body angle. We also tested for an effect of ICP in the pulsatility model, given pulsatility is expected to increase with ICP.^36^ Models included random effects of participant, to account for clustering of data within an individual, and head position, to allow slopes to vary by head position. We found minimal differences between seated and standing (i.e. upright) body positions (Fig. 1, Fig. 4A); likelihood ratio testing also confirmed no benefit to model fit when body position included both seated and standing positions rather than simply upright body position. We therefore pooled sitting and standing to upright body positions and compared with supine body position. Model assumptions were assessed by visual inspection of residuals. We report two-sided p values (alpha level: p<0.05), Wald tests of contrasts, estimated marginal means and 95% confidence intervals. Post-hoc tests were adjusted for multiplicity using the Šidák method.

## RESULTS

### ICP and pulsatility in head neutral position

We found no evidence that ICP or pulsatility in the head neutral position varied by shunt or diagnosed pathology (all p>0.05). As expected, ICP varied by body position (χ^2^=487.2, p<0.001). ICP was greater in supine than seated or standing positions (p<0.001) but did not differ between seated and standing positions (p=0.997; supine: 12.4 mmHg [10.7,14.2], seated: 1.7 mmHg [0.2,3.2], standing: 1.8 mmHg [0.2,3.4]). Pulsatility also varied by body position (χ^2^=12.0, p=0.003) but the differences were more subtle than with ICP. Pulsatility was greater in seated than supine body position (p=0.002) but did not differ between seated and standing (p=0.065) or standing and supine body positions (p=0.681; supine: 4.1 mmHg [3.6,4.6], seated: 4.9 mmHg [4.4,5.4], standing: 4.3 mmHg [3.8,4.9]). The lack of difference in ICP and pulsatility between seated and standing positions further justified reducing to upright positions.

### Effect of head turn and forward tilt on ICP in upright and supine body positions

Fig. 1 and Fig. 2 show the change in ICP when tilting the head forward or turning the head right or left for 20s in upright (seated, standing) and supine body positions. ICP increased on average when turning the head right or left in upright and supine body positions and when tilting the head forward in upright body positions (Fig. 1). The increase in ICP tended to occur at the same time the head was moved into position and returned to the prior baseline after the head was moved back to the neutral position. The elevated ICP was relatively sustained over the 20s period that the head position was adopted. However, little to no change in ICP occurred on average when tilting the head forward when supine (Fig. 1 bottom right).

**Figure 2.**
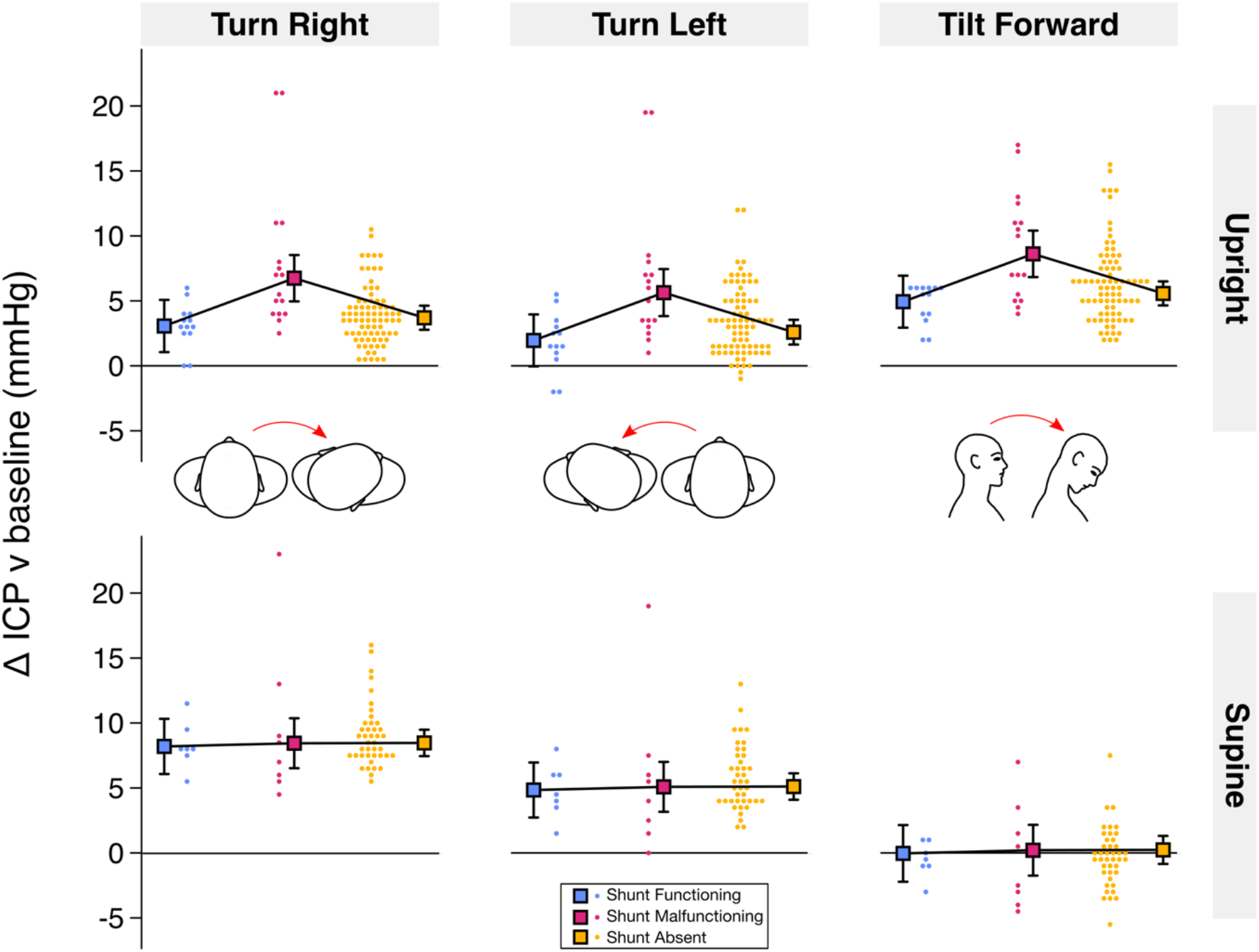
Effect of head turn and forward tilt on ICP. Change in ICP relative to baseline (head neutral) averaged over 20s in each head position (left column: turn right; middle column: turn left; right column: tilt forward), body position (top row: upright [seated, standing]; bottom row: supine) and shunt status group (blue: shunt functioning; magenta: shunt malfunctioning; yellow: shunt absent). Squares with error bars are estimated marginal means with 95% CIs and circles are fitted values for each individual from mixed effects modelling.

Statistical modelling confirmed these observations (Fig. 2). The ICP model of best fit included the terms head-on-body angle, a two-way shunt by body position interaction, and a three-way head position by body position by Postural Orthostatic Tachycardia Syndrome (POTS) interaction. ICP increased significantly relative to baseline in each head position (p<0.001; Table 2), except for forward tilt in supine body position where ICP did not change on average (forward tilt-supine v baseline: 0.3 mmHg [-0.8,1.3], p=0.596; head by body position: χ^2^=184.1, p<0.001; Fig. 2; Table 2).

**Table 2.** Change in ICP and pulsatility versus head neutral in each head and body position.

|  | Upright body position |  |  |  |  |  | Supine body position |  |  |
| --- | --- | --- | --- | --- | --- | --- | --- | --- | --- |
|  | Turn |  | Tilt |  | Lateral Tilt |  | Turn |  | Tilt |
|  | Right | Left | Forward | Backward | Right | Left | Right | Left | Forward |
| ICP (mmHg) | 4.4 [3.5,5.3] | 3.2 [2.3,4.1] | 6.4 [5.5,7.3] | 7.4 [5.0,9.8] | 4.3 [3.5,5.0] | 2.7 [1.9,3.4] | 9.1 [8.1,10.1] | 5.5 [4.5,6.5] | 0.3 [-0.8,1.3] |
| Pulsatility (mmHg) | 1.6 [1.3,1.9] | 1.9 [1.6,2.2] | -0.7 [-1.0,-0.4] | 2.7 [1.4,4.1] | 0.3 [0.2,0.5] | 0.7 [0.5,0.9] | 1.8 [1.4,2.1] | 2.0 [1.7,2.3] | 1.5 [1.1,1.9] |

Note: Upright head neutral ICP=1.7 mmHg [0.3,3.2]; supine head neutral ICP=12.4 mmHg [10.7,14.2]; upright head neutral pulsatility=4.6 mmHg [4.2,5.0]; supine head neutral pulsatility=4.1 mmHg [3.6,4.6]. Values are estimated marginal means and 95% CIs from mixed effects models.

ICP increased more with right than left head turn in supine (right-left difference: 4.0 mmHg [0.5,7.5]; p=0.010) but not upright body positions (right-left difference: 0.4 mmHg [– 2.5,3.2]; p>0.999). The increase in ICP with right head turn was greater in supine than upright body positions (supine-upright difference: 5.0 mmHg [2.5,7.6]; p<0.001) but did not differ by body position in left head turn (supine-upright difference: 0.7 mmHg [-1.9,3.3]; p>0.999). Head tilt forward increased ICP more than head turn right or left in upright body positions (tilt forward-turn right difference: 4.3 mmHg [1.5,7.2]; p<0.001; tilt forward-turn left difference: 4.0 mmHg [1.1,6.8]; p=0.001). The increase in ICP with forward tilt in upright body positions was particularly great in the 3 patients with POTS (POTS: 12.8 mmHg [9.0,16.7], no POTS: 6.1 mmHg [5.1,7.0]; p=0.005). No differences were observed with POTS in other head and body positions (all p>0.07).

There was evidence of an effect of shunt in upright (χ^2^=11.1, p=0.004) but not supine body positions (χ^2^=0.06, p=0.972; Fig. 2). The increase in ICP with head turn and forward tilt in upright body positions was greater in patients with shunt malfunction than with a functioning shunt and no shunt (p<0.001; shunt malfunction: 7.6 mmHg [5.7,9.4], shunt function: 3.6 mmHg [1.5,5.7], no shunt: 4.3 mmHg [3.4,5.2]; Fig. 2). The increase in ICP tended to be smaller in patients with a functioning shunt than with no shunt (Fig. 1) but this was not statistically significant either in upright body positions (p=0.912) or across head and body positions (shunt: χ^2^=3.0, p=0.223).

A greater increase in ICP was associated with greater head-on-body angle (β=0.039 mmHg/degrees [0.015,0.063], p=0.002; predicted 1 mmHg increase per 25 degrees).

Baseline ICP, diagnosed pathologies other than POTS (including IIH or Chiari malformation), age, sex, BMI, and hypertension did not improve model fit implying no consistent effect on ICP in head turn or forward tilt positions.

### Effect of head turn and forward tilt on pulsatility in upright and supine body positions

Fig. 3 shows the change in pulsatility when tilting the head forward or turning the head right or left for 20s in upright (seated, standing) and supine body positions. The pulsatility model of best fit included a three-way head position by body position by POTS interaction and the change in ICP from baseline. After adjustment for the change in ICP, pulsatility increased significantly relative to baseline in each head position (p<0.001; Table 2), except for forward tilt in upright body positions where pulsatility decreased on average (forward tilt-upright v baseline: –0.7 mmHg [-1.0,-0.4], p<0.001; head by body position: χ^2^=6.6, p=0.036; Fig. 3; Table 2). The increase in pulsatility did not differ between left and right head turn in either upright or supine body positions and forward tilt when supine (all p>0.999). On average pulsatility increased by 1.5-2 mmHg (Table 2). No differences in pulsatility were observed with POTS in any combination of head and body position (all p>0.247). Pulsatility increased by 0.2 mmHg for every 1 mmHg increase in ICP from baseline when the head was tilted forward or turned to the side (β=0.199 mmHg/mmHg [0.169,0.229], p<0.001).

**Figure 3.**
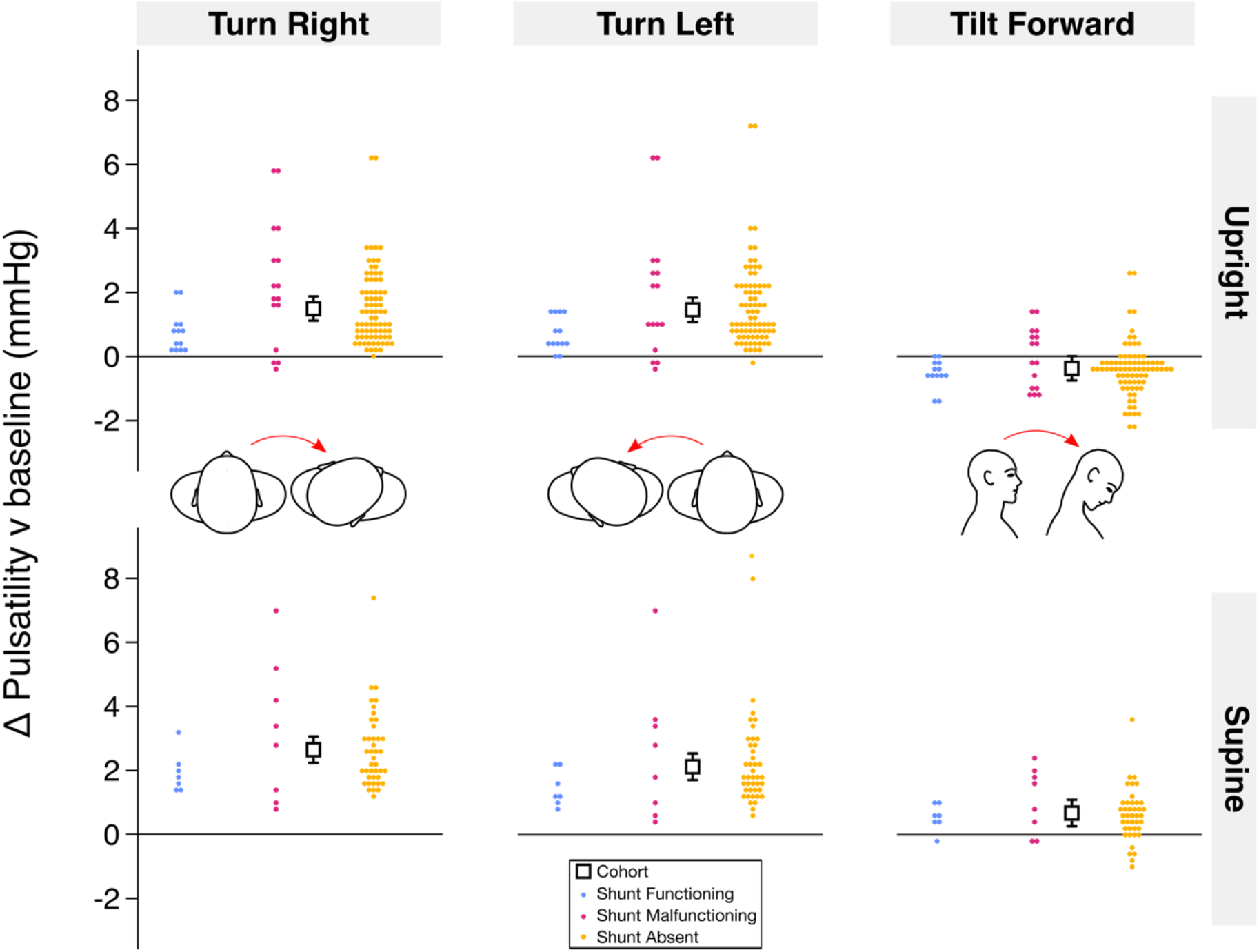
Effect of head turn and forward tilt on pulsatility. Change in pulsatility relative to baseline (head neutral) averaged over 20s in each head position (left column: turn right; middle column: turn left; right column: tilt forward), body position (top row: upright [seated, standing]; bottom row: supine) and shunt status group (blue: shunt functioning; magenta: shunt malfunctioning; yellow: shunt absent). Squares with error bars are estimated marginal means with 95% CIs for the whole cohort and circles are fitted values for each individual from mixed effects modelling.

Baseline ICP, baseline pulsatility, shunt status, diagnosed pathologies other than POTS (including IIH or Chiari malformation), age, sex, BMI, head-on-body angle, and hypertension did not improve model fit implying no consistent effect on pulsatility in head turn or forward tilt positions.

### Effect of head lateral tilt on ICP and pulsatility in upright body positions

Fig. 4A shows the change in ICP and pulsatility when tilting the head laterally to the right or left for 20s in upright (seated, standing) body positions. Similar to turning the head to the side or tilting the head forward, lateral head tilt increased ICP and pulsatility (Fig. 4B, Fig. 4C).

**Figure 4.**
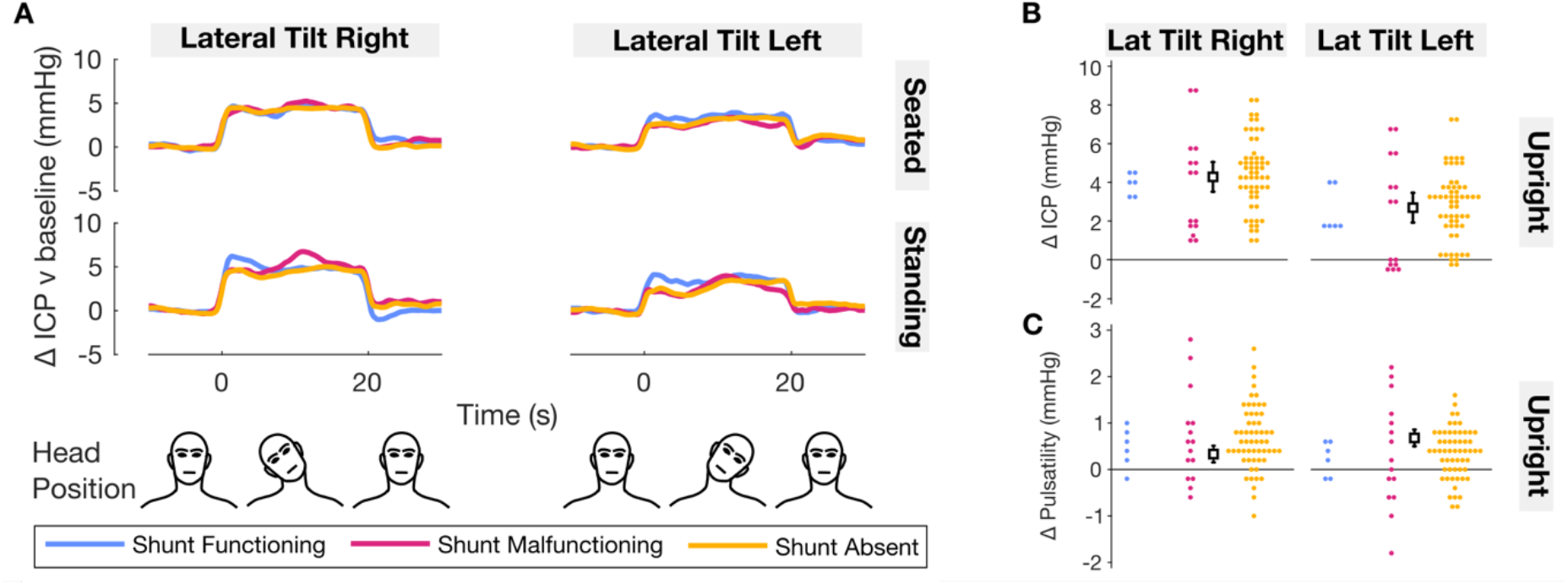
Effect of head lateral tilt on ICP and pulsatility. A: Traces are the mean change in ICP over time relative to baseline (head neutral) for each head position (left column: lateral tilt right; right column: lateral tilt left) in each body position (top row: seated; bottom row: standing) and shunt status group (blue: shunt functioning; magenta: shunt malfunctioning; yellow: shunt absent). The head was moved from neutral into each position at time=0s and back to neutral at time=20s. For plotting, ICP was averaged from pulse-to-pulse to remove pulsatile variation prior to averaging per individual and group. Graphics at bottom of figure aid interpretation of head positions and timings. B, C: Change in ICP (B) and pulsatility (C) relative to baseline (head neutral) averaged over 20s in each head position (left column: lateral tilt right; right column: lateral tilt left) in upright (seated, standing) body positions (bottom row) and shunt status group. Squares with error bars are estimated marginal means with 95% CIs for the whole cohort and circles are fitted values for each individual from mixed effects modelling.

Separate statistical models were used to analyse lateral tilt data (see ‘Statistical analysis’). The ICP model of best fit included the term head position only. ICP increased by 3.5 mmHg [2.8,4.2] on average but the increase was greater by 1.6 mmHg [0.9,2.2] in right than left lateral tilt (head position: p<0.001; Fig. 4B; Table 2).

The pulsatility model of best fit included the change in ICP from baseline only. Pulsatility increased by 0.2 mmHg for every 1 mmHg increase in ICP from baseline when the head was tilted laterally (β=0.203 mmHg/mmHg [0.164,0.242], p<0.001). After adjustment for the change in ICP, pulsatility increased significantly relative to baseline by 0.5 mmHg on average ([0.3,0.7]; lateral tilt v baseline: p<0.001; Fig. 4C; Table 2).

Baseline ICP, baseline pulsatility, shunt status, diagnosed pathology, age, sex, BMI, head-on-body angle, and hypertension did not improve model fit implying no consistent effect on ICP or pulsatility in head lateral tilt position.

### Effect of head backward tilt on ICP and pulsatility in seated body position

Fig. 5A shows the change in ICP and pulsatility when tilting the head backward for 10s in seated body position. Similar to tilting the head forwards whilst seated, backwards tilt increased ICP. Separate statistical models were used to analyse backward tilt data (see ‘Statistical analysis’). No tested terms improved ICP model fit, even when using a less strict alpha level (all p>0.05). ICP increased by 7.4 mmHg [5.0,9.8] relative to baseline (p<0.001) during backward tilt (Fig. 5B; Table 2) and did not differ from the first 10s of forward tilt (p=0.845).

**Figure 5.**
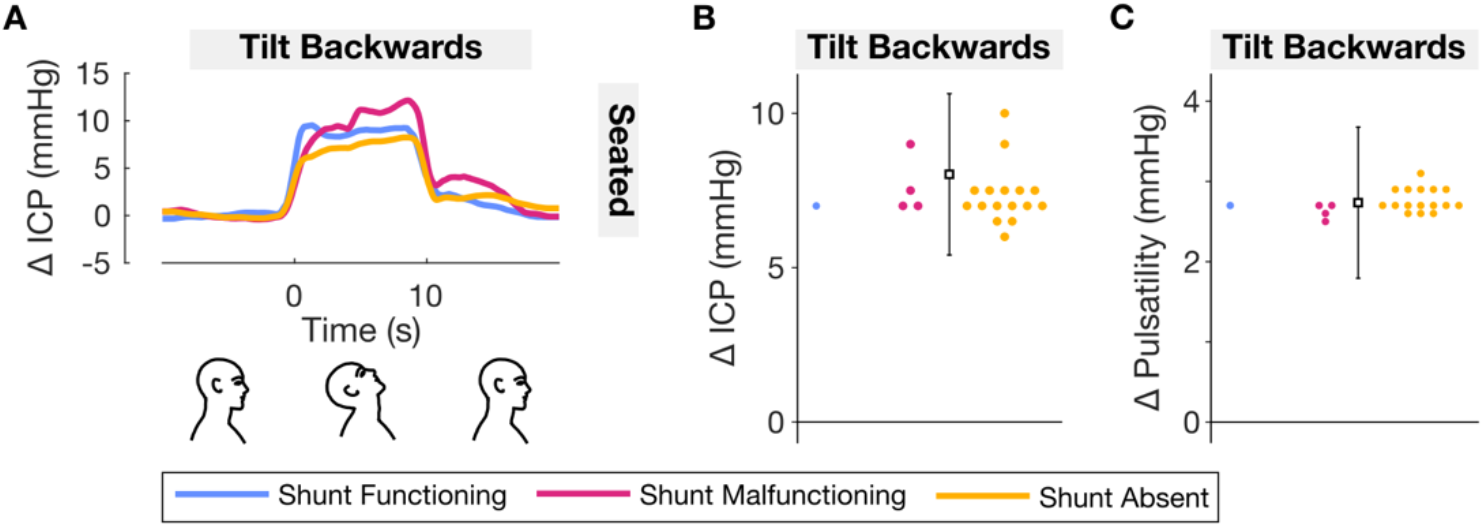
Effect of head backward tilt on ICP and pulsatility. A: Traces are the mean change in ICP over time relative to baseline (head neutral) for head backward tilt in seated body position for each shunt status group (blue: shunt functioning; magenta: shunt malfunctioning; yellow: shunt absent). The head was moved from neutral into position at time=0s and back to neutral at time=10s. For plotting, ICP was averaged from pulse-to-pulse to remove pulsatile variation prior to averaging per individual and group. Graphics at bottom of figure aid interpretation of head positions and timings. B, C: Change in ICP (B) and pulsatility (C) relative to baseline (head neutral) averaged over 10s in head backward tilt position (left column: lateral tilt right; right column: lateral tilt left) when seated for each shunt status group. Squares with error bars are estimated marginal means with 95% CIs for the whole cohort and circles are fitted values for each individual from mixed effects modelling.

In contrast to forward tilt in seated body position where pulsatility decreased, backwards tilt increased pulsatility (Fig. 5C). No tested terms improved pulsatility model fit. Pulsatility increased by 2.7 mmHg [1.4,4.1] relative to baseline during backward tilt (p<0.001; Table 2).

## DISCUSSION

We measured parenchymal ICP and pulsatility in different combinations of head and body positions in a relatively well, ambulatory cohort of mixed diagnosed pathology and shunt status. ICP and pulsatility tended to increase when the head was positioned away from the neutral position. Notable exceptions were head forward tilt in supine body position, which increased pulsatility but did not change ICP, and head forward tilt in upright body positions, which increased ICP but decreased pulsatility. The increase in ICP with head turn and forward tilt in upright body positions was greatest in individuals with a malfunctioning shunt. Together, this demonstrates that: (1) the position of the head on the body influences ICP and pulsatility; (2) the effect of the head-on-body position on ICP and pulsatility depends on body position; and (3) shunt status can influence the effect of head-on-body position on ICP.

### ICP and pulsatility depend on a combination of head and body position

Moving the head away from the neutral position did not have a constant effect on ICP and pulsatility; instead, the effect depended on a combination of head and body position. When in upright body positions, ICP increased when the head was turned or tilted to either side or when tilted forward or backward (Figs. 1-5; Table 2). When in supine body position, ICP also increased when the head was turned to either side but did not change when tilted forward (Figs. 1-2; Table 2). The lack of change in ICP with head forward tilt whilst supine was notable as the only position tested where ICP did not increase relative to the head neutral position.

Moving the head away from neutral into each head position compresses neck vasculature and presumably reduced venous outflow from the skull to contribute to the increase in ICP. ^19,24–30^ Head turn of similar magnitude to that observed here (Supplementary Fig. 1) occludes the internal jugular veins in 80% of individuals.^30^ In upright body positions, head tilt forwards, backwards or laterally moved the head from upright towards horizontal (Supplementary Fig. 1). This presumably also contributed to the increase in ICP via gravitational hydrostatic effects and/or displacement of the brain within the skull. However, head forward tilt whilst supine moved the head in the reverse direction, i.e. from horizontal towards an upright position, which presumably decreased ICP. The neck vasculature-induced increase may have counteracted the gravitational-induced decrease to result in no change to ICP on average in head forward tilt whilst supine.

ICP increased more when the head was moved to the right than left with lateral tilt in upright body positions and head turn in supine body positions.

The greater increase in ICP in right than left lateral tilt in upright body positions and head turn in supine body position is likely due to the aforementioned gravitational effects of the predominantly right-sided ICP probe placement in our patients, but may also be due to the higher prevalence of right-than left-dominant transverse venous sinus drainage.^24^

Pulsatility increased by ∼0.5-2 mmHg on average when the head was turned or tilted to the side in upright or supine body positions, and when the head was tilted forwards when supine (Table 2). This represented a 12.5-50% increase in pulsatility relative to the head neutral position and implies cranial compliance decreased in each of these head positions, even though head position was maintained for a relatively brief duration (up to 20 s).

### Shunt

The influence of head position on ICP and pulsatility was similar across shunt status groups, i.e. ICP and pulsatility increased in each group in most combinations of head and body position. However, the increase in ICP with head turn and forward tilt in upright body positions was larger in individuals with a malfunctioning shunt than with either a functioning shunt or no shunt. Shunted individuals may be more dependent on their shunt for ICP control in upright body positions, where the physiological collapse of internal jugular veins partially blocks an alternative venous outflow pathway.^46,47^ A malfunctioning shunt may have prevented relief of excess pressure that otherwise would have been possible if the shunt was functioning. It is unclear why no differences between shunt groups were observed when supine, but re-opening of the IJVs when supine may have allowed sufficient outflow of intracranial fluid to prevent excess pressure when the head was positioned away from neutral.

### IIH, Chiari malformation and other pathologies

Our cohort contained a range of diagnosed pathologies, including IIH, Chiari malformation, syrinx, Ehlers-Danlos syndrome, and POTS. In the absence of prior data to inform specific hypotheses, we tested whether each pathology and its interaction with head and body position improved ICP and pulsatility model fit, but with stringent statistical criteria to mitigate the false positive rate. POTS was the only pathology to improve model fit. ICP increased more in those with than without POTS when the head was tilted forward in an upright body position. Future work should determine whether this finding generalises beyond the three individuals with POTS in our cohort. Whilst the lack of improvement in model fit with IIH, Chiari malformation, syrinx, and Ehlers-Danlos syndrome implies no consistent influence of each pathology on ICP and pulsatility in different head positions, we may be underpowered to detect any difference with n<13 in each pathology. However, the improvement to model fit generally did not approach significance in pathologies other than POTS, even when using less stringent statistical criteria.

### Clinical importance

Patients with abnormal ICP dynamics often report symptoms induced and/or exacerbated by head movement and so avoid excess head movement. For example, a preferred strategy to look to the side might involve turning the body whilst maintaining a neutral head position instead of turning the head. Our finding that ICP and pulsatility generally increase when the head is positioned away from neutral provides potential reason for this behaviour.

Our findings also highlight the importance of standardising head position when ICP is measured clinically. The increase in ICP was 3-9 mmHg on average (Table 2) with the head moved away from neutral and could be large enough to affect treatment decisions.^28^ Whilst it is unlikely ICP would be measured with the head moved as far from neutral as in our study, it is important to note that the magnitude of increase varied between individuals and increased more the further the head was moved from neutral. Clinicians should be cautious that even relatively small movements of the head away from neutral could influence ICP in some individuals. Future work should establish the clinical utility of measuring ICP and pulsatility in different head positions, for example as a biomarker of shunt malfunction.^48^

Future treatments such as the proposed ‘smart’ CSF-diverting shunt^8^ depend on improved understanding of ICP dynamics in activities of daily living. Our findings highlight that future smart shunt design should account for ICP changes in different head positions, not only in different body positions. Our data describes the predicted change in ICP in different head and body positions and could underpin future smart shunt design.

## DATA AVAILABILITY

The data that support the findings of this study are available from the authors upon reasonable request.

## ACKNOWLEDGMENTS

We thank the patients who participated in the study for their time and efforts.

## FUNDING

The project was supported by an innovation fund grant from the National Brain Appeal. EMM was supported by a research fellowship sponsored by B.Braun. AKT’s research time was supported by the National Institute for Health Research University College London Hospitals Biomedical Research Centre.

## COMPETING INTERESTS

EMM was supported by a research fellowship sponsored by B.Braun. LDW has received honoraria from and served on advisory boards for Medtronic, B. Braun and Codman.

## SUPPLEMENTARY MATERIALS

**Supplementary Figure 1.**
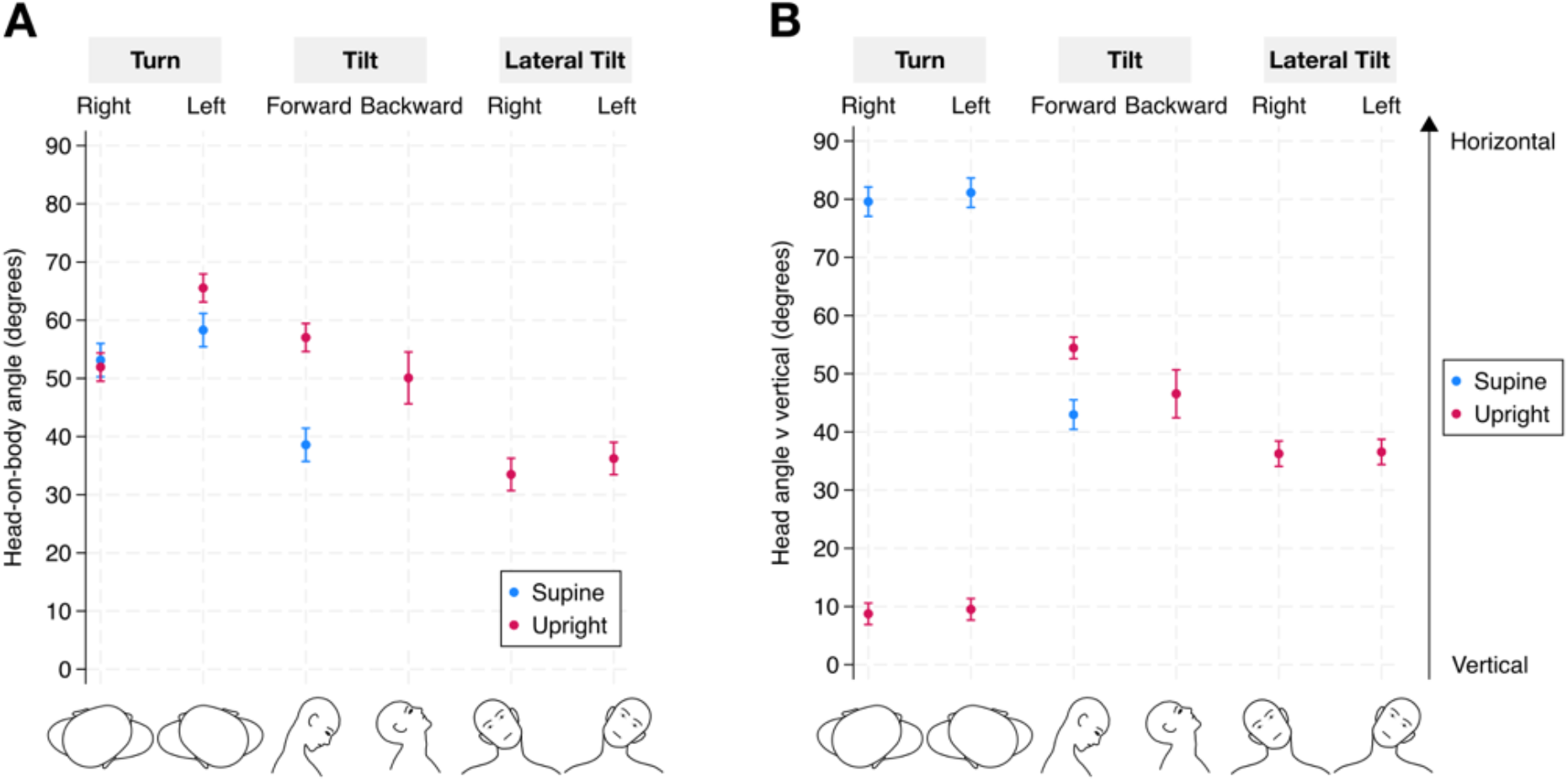
Head-on-body angle and head angle versus vertical in each head and body position. A: Head-on-body angle in each head position for each body position. Head-on-body angle was computed as the change in head relative to body orientation from head neutral position (see Methods). B: Head angle relative to gravitational vertical. Note the head was approximately aligned with gravitational vertical in head neutral-upright body positions but approximately 90 degrees relative to gravitational vertical in head neutral-supine body position. Head-on-body angle approximated the head angle versus vertical for head positions involving a predominant movement of the head in the vertical plane (tilt, lateral tilt). Blue=supine; pink=upright (seated, standing). Filled circles are estimated marginal means from mixed effects modelling; error bars are 95% confidence intervals. Graphics below A and B aid interpretation of each head position.

**Supplementary Table 1.** Change in head-on-body angle versus head neutral and head angle versus vertical in each head and body position.

|  | Upright body position |  |  |  |  |  | Supine body position |  |  |
| --- | --- | --- | --- | --- | --- | --- | --- | --- | --- |
|  | Turn |  | Tilt |  | Lateral Tilt |  | Turn |  | Tilt |
|  | Right | Left | Forward | Backward | Right | Left | Right | Left | Forward |
| Head-on-body angle (deg) | 52 | 66 | 57 | 50 | 33 | 36 | 53 | 58 | 38 |
|  | [50,54] | [63,68] | [55,59] | [46,55] | [31,36] | [33,39] | [50,56] | [55,61] | [36,41] |
| Head verticality (deg) | 9 | 10 | 54 | 47 | 36 | 36 | 80 | 81 | 43 |
|  | [8,11] | [8,11] | [52,56] | [42,51] | [34,38] | [34,39] | [77,82] | [79,84] | [40,46] |

Note: Values are estimated marginal means and 95% CIs from mixed effects models.

## Notes

### Author Declarations

The study conformed to the Declaration of Helsinki and ethical approval was granted by local Research Ethics Committee (UCLH project ID 15/0769).

